# A randomized controlled trial of inhaled ciclesonide for outpatient treatment of symptomatic COVID-19 infections

**DOI:** 10.1101/2021.09.07.21261811

**Authors:** Brian M. Clemency, Renoj Varughese, Yaneicy Gonzalez-Rojas, Caryn G. Morse, Wanda Phipatanakul, David J. Koster, Michael S. Blaiss

## Abstract

**Importance:** Systemic corticosteroids are commonly used in the treatment of severe COVID-19. However, their role in the treatment of patients with mild to moderate disease is less clear. The inhaled corticosteroid ciclesonide has shown early promise as a potential treatment for COVID-19.

**Objective:** To determine whether the inhaled steroid ciclesonide is efficacious in patients with high risk for disease progression and can reduce the incidence of long-term COVID-19 symptoms or post-acute sequelae of SARS-CoV-2.

**Design:** This was a phase III, multicenter, double-blind, randomized controlled trial to assess the safety and efficacy of ciclesonide metered-dose inhaler (MDI) for the treatment of non-hospitalized participants with symptomatic COVID-19 infection. Patients were screened from June 11, 2020 to November 3, 2020.

**Setting:** The study was conducted at 10 centers throughout the U.S. public and private, academic and non-academic sites were represented among the centers.

**Participants:** Participants were randomly assigned to ciclesonide MDI 160 µg per actuation, two actuations twice a day (total daily dose 640 µg) or placebo for 30 days.

**Main Outcomes and Measures:** The primary endpoint was time to alleviation of all COVID-19 related symptoms (cough, dyspnea, chills, feeling feverish, repeated shaking with chills, muscle pain, headache, sore throat, and new loss of taste or smell) by Day 30. Secondary endpoints included subsequent emergency department visits or hospital admissions for reasons attributable to COVID-19.

**Results:** 413 participants were screened and 400 (96.9%) were enrolled and randomized (197 in the ciclesonide arm and 203 in the placebo arm). The median time to alleviation of all COVID-19-related symptoms was 19.0 days (95% CI: 14.0, 21.0) in the ciclesonide arm and 19.0 days (95% CI: 16.0, 23.0) in the placebo arm. There was no difference in resolution of all symptoms by Day 30 (odds ratio [OR] 1.28, 95% CI: 0.84, 1.97). Participants treated with ciclesonide had fewer subsequent emergency department visits or hospital admissions for reasons attributable to COVID-19 (OR 0.18, 95% CI: 0.04 - 0.85). No subjects died during the study.

**Conclusions and Relevance:** Ciclesonide did not achieve the primary efficacy endpoint of time to alleviation of all COVID-19-related symptoms. Future studies of inhaled steroids are needed to explore their efficacy in patients with high risk for disease progression and in reducing the incidence of long-term COVID-19 symptoms or post-acute sequelae of SARS-CoV-2.

**Trial Registration:** ClinicalTrials.gov

NCT04377711

https://clinicaltrials.gov/ct2/show/NCT04377711

**Key Points:** *Question:* Can the inhaled steroid ciclesonide be efficacious in patients with high risk for disease progression and reduce the incidence of long-term COVID-19 symptoms or post-acute sequelae of SARS-CoV-2?

*Findings:* In this randomized clinical trial of 413 patients, ciclesonide did not reduce the time to alleviation of all COVID-19-related symptoms. However, patients treated with ciclesonide had fewer subsequent emergency department visits or hospital admissions for reasons attributable to COVID-19.

*Meaning:* Future studies of inhaled steroids are needed to explore their efficacy in patients with high risk for disease progression and in reducing the incidence of long-term COVID-19 symptoms or post-acute sequelae of SARS-CoV-2.

## Introduction

The SARS-CoV-2 coronavirus disease (COVID-19) pandemic has led to an ongoing global public health emergency. Symptoms of COVID-19 vary widely and include fever, cough, difficulty breathing, and loss of taste and smell. Reported illnesses have ranged from asymptomatic to severe illness and death from confirmed COVID-19 cases. The antiviral, remdesivir, was the first agent to receive approval from the United States Food and Drug Administration (FDA) for the treatment of COVID-19^1^. To date, the majority of therapeutic studies have focused on patients with severe disease requiring hospitalization.

In the RECOVERY trial, dexamethasone resulted in lower 28-day mortality among patients with severe COVID-19^2^. The role of corticosteroids for patients with mild to moderate coronavirus disease is less clear, as systemic corticosteroids have shown mixed results for these patient types^3^. The potential anti-inflammatory benefits of corticosteroids must be weighed against the potential risks of immunosuppression and other systemic steroid effects^4,5^.

Inhaled corticosteroids may also be beneficial in COVID-19 treatment, as they reduce the expression of key proteins involved in the entry of the virus into host cells^6^. Inhaled corticosteroids have also been showed to cause downregulation of COVID-19 genes^7^.

Among the available inhaled corticosteroids, ciclesonide has emerged as a potential treatment option for COVID-19. In vitro, ciclesonide has been shown to have antiviral properties against COVID-19 and blocks COVID-19 viral replication^8,9^. A case series described three elderly patients with hypoxia due to COVID-19 who recovered following treatment with ciclesonide^10^. Clinical trials are needed to determine the effects of ciclesonide on COVID-19 in the clinical setting. This study examined the effects of ciclesonide versus placebo in non-hospitalized participants with symptomatic COVID-19 infection.

## Methods

### Study design and participants

This was a phase III, multicenter, double-blinded, randomized placebo-controlled trial to assess the safety and efficacy of ciclesonide metered-dose inhaler (MDI) for the treatment of non-hospitalized patients with symptomatic COVID-19 infection. Participants were eligible for inclusion if, at the time of enrollment, they (1) were at least 12 years of age, (2) had a positive SARS-CoV-2 molecular or antigen diagnostic sample obtained in the previous 72 hours, (3) were not hospitalized or under consideration for hospitalization, (4) had an oxygen saturation of at least 93% on room air, (5) were able to demonstrate successful use of an MDI, and (6) had at least one of the following symptoms of COVID: fever, cough, or dyspnea. Twelve years of age was selected as the age threshold for enrollment, consistent with the FDA approved prescribing information for ciclesonide for the maintenance treatment of asthma as prophylactic therapy. Participants were excluded if they (1) had a history of hypersensitivity to ciclesonide, (2) had taken an inhaled or intranasal corticosteroid within 14 days, (3) had taken oral corticosteroids within 90 days, (4) had participated in any other clinical trial or use of any investigational agent within 30 days, (5) had a history of cystic fibrosis, (6) has a history of idiopathic pulmonary fibrosis, (7) were receiving treatment with hydroxychloroquine/chloroquine, or (8) were pregnant.

The study was conducted at 10 centers throughout the U.S. Public and private, academic and non-academic sites were represented among the centers. This study was approved by the Western Institutional Review Board (IRB). All participants, parents/legal guardians, or legally authorized representatives provided written informed consent.

### Randomization and Masking

Approximately 400 eligible participants were planned to be randomized 1:1 to receive treatment with ciclesonide MDI 160 µg per actuation, two actuations (AM and PM) twice a day (total daily dose 640 µg), plus standard supportive care or placebo MDI twice a day (BID) plus standard supportive care for 30 days. Standard supportive care was provided at the discretion of the study nurse or physician and included recommendations for symptomatic medications and directions to seek emergency care when necessary. The total daily dose of 640 µg was selected for this study, consistent with the highest recommend daily dose in the FDA approved prescribing information for ciclesonide for the maintenance treatment of asthma as prophylactic therapy.

Ciclesonide MDIs and placebo MDIs were identical in appearance. The randomization schedule was generated by the contract manufacturing organization and incorporated into the labeling of kits. MDI kits were sent to the study sites in blocks of 6 with 3 active and 3 placebo kits randomized within each block. Individual site personnel dispensed individual kits in order, blinded to the assignment.

### Procedures

Participants were instructed on how to self-administer the MDI at the initial visit by the study team. MDI use was reviewed with participants during all follow-up calls. MDI technique was not evaluated after the initial visits. Participants were dispensed a 30-day supply of investigational product, a pulse oximeter for at-home oxygen saturation level monitoring, and an electronic diary smartphone application (eDiary). Within one hour of self-administration of the investigational product, participants were to complete and record in their eDiary the presence of COVID-19-related symptoms of cough, dyspnea, chills, feeling feverish, repeated shaking with chills, muscle pain, headache, sore throat, and new loss of taste or smell. Participants received reminders to self-administer the study medication and log symptoms in the form of push notifications from their eDiary and scheduled phone calls from the study team. Qualified healthcare providers contacted participants on Days 2, 4, 6, 8, 10, 12, 14, 21 ± 2 days and conducted a study visit on Day 30 ± 2 days for a health status check to collect adverse event and concomitant medication information, and to confirm and/or clarify information recorded in the eDiary. The healthcare providers also contacted participants on Day 37 ± 4 days and Day 60 ± 7 days to collect follow-up safety and outcome data. A nasopharyngeal sample for quantitative viral load analysis was obtained at the initial visit and on Day 30 ± 2 to correspond with the start and end of the treatment protocol.

Per the protocol, participants were instructed to seek emergency department evaluation if their oxygen saturation was less than or equal to 92%. Participants (or their representatives) were asked to notify study personnel directly in the event they visited an emergency department or were hospitalized during their participation in the study. Participants were instructed to continue the study medication for 30 days, even if symptoms resolved earlier. Continuing the medication for 30 days standardized administrations schemes for all patients and allowed for the evaluation of symptom based and non-symptom-based outcome measures.

### Outcomes

The primary outcome was time to alleviation of all COVID-19 related symptoms (cough, dyspnea, chills, feeling feverish, repeated shaking with chills, muscle pain, headache, sore throat, and new loss of taste or smell) by Day 30. The time to alleviation of COVID-19-related symptoms was defined as being symptom-free for a continuous period of at least 24 hours (i.e., at least 3 consecutive AM/PM assessments), as self-reported in the participant’s eDiary.

Secondary outcomes were to assess whether ciclesonide MDI plus standard supportive care reduces the incidence of subsequent emergency department visits or hospital admissions for reasons attributable to COVID-19, reduces the incidence of hospital admissions or death, reduces all-cause mortality, reduces COVID-19-related mortality, increases the percentage of participants with alleviation of COVID-19-related symptoms and increases the time to hospital admission or death compared with placebo plus standard supportive care. Alleviation of all COVID-19 related symptoms by Days 7, 14, and 30 were also compared. Additional secondary outcomes included oxygen saturation levels, COVID-19 viral load, and safety assessments.

### Statistical Analysis

The primary efficacy analysis was based on the intent-to-treat (ITT) population (all randomized participants). A sensitivity analysis was performed for the per-protocol (PP) population. Participant eDiary compliance of < 65% was considered a major protocol deviation, as a proxy for medication non-compliance. To improve treatment-effect estimation and inference precision, preplanned baseline covariate adjustments were made for sex, age, race, and body mass index (BMI) as these are known COVID-19 risk factors. A Cox proportional hazard model was used to allow for the inclusion of these additional covariates. The median time to event and 95% confidence intervals (CIs) were summarized by treatment arm, and Kaplan-Meier estimates of the survival curves were generated. A shift table depicting the change in severity of COVID-19-related symptoms of cough, dyspnea, chills, and feeling feverish from baseline was presented for each day and timepoint.

The secondary efficacy analysis was based on the ITT population. A sensitivity analysis was performed for the PP population. The secondary efficacy endpoints were analyzed using a logistic regression model, except for viral load and oxygen saturation level which were analyzed using an analysis of covariance (ANCOVA) model.

The ITT population was used to evaluate safety. All adverse events and serious adverse events were coded using Medical Dictionary for Regulatory Activities v23.0. The Data Monitoring Committee conducted a review of blinded safety data once 100 participants had been enrolled.

### Primary Outcome and Sample Size Changes

The original primary outcome registered was percentage of participants with subsequent emergency department visit or hospital admission for reasons attributable to COVID-19 by Day The enrollment target of 400 subjects was based on sample size calculations for this original end point.

The primary endpoint was changed in subsequent versions of the protocol. The final primary outcome, which is the primary outcome reported in this manuscript, was time to alleviation of all COVID-19-related symptoms by Day 30. A primary outcome based on symptom resolution was chosen rather than one based on emergency department visits or hospital admission after preliminary data demonstrated substantially lower than expected rates of emergency department visits or hospitalizations among study participants. For the final primary outcome, a sample size of approximately 232 patients (116 per arm) was required to achieve 90% power at α = 0.05.

This was based on the assumptions that there would be a median time to alleviation of symptoms of 7 days for the ciclesonide arm and 11 days for the placebo arm (hazard ratio of approximately 1.58) with a total study duration of 30 days. The larger enrollment target of 400 subjects was maintained.

This trial was prospectively registered with ClinicalTrials.gov, NCT04377711. The changes made to the primary outcome were incorporated into the protocol, approved by the IRB, were reflected in updates to the study registration and had no effect on the data collection process. The final version of the protocol will be available on ClinicalTrials.gov.

## Results

From June 11, 2020 to November 3, 2020, 413 participants were screened and 400 (96.9%) were enrolled and randomized (197 in the ciclesonide arm and 203 in the placebo arm). All randomized participants received at least one dose of investigational product and were included in both the ITT and the safety populations. The PP population included 377 (94.3%) participants (184 [93.4%] participants in the ciclesonide arm and 193 [95.1%] participants in the placebo arm). One subject in the ciclesonide arm was excluded from the PP due to a pregnancy test being performed prior to signing the informed consent, the remaining 22 participants (12 in the ciclesonide arm, 10 in the placebo arm) were excluded from the PP due to diary compliance < 65%.

Overall, 359 (89.8%) participants (178 [90.4%] participants in the ciclesonide arm and 181 [89.2%] participants in the placebo arm) completed the study and 41 (10.3%) participants (19 [9.6%] and 22 [10.8%], respectively) discontinued. For participants in both arms, the most common reason for discontinuation was that participants were lost to follow-up (11 [5.6%] participants in the ciclesonide arm and 9 [4.4%] participants in the placebo arm) (Figure 1). In the ITT population, 55.3% were female, 11.8% were black or African American, and 43.0% were Hispanic or Latino. The ciclesonide arm had higher rates of Type 2 diabetes mellitus (p=0.007) and asthma p=0.042); otherwise, demographic characteristics, medical histories and concomitant medications of interest were not different between the arms (Table 1). No participants were treated with remdesivir during the study.

**Table 1.**
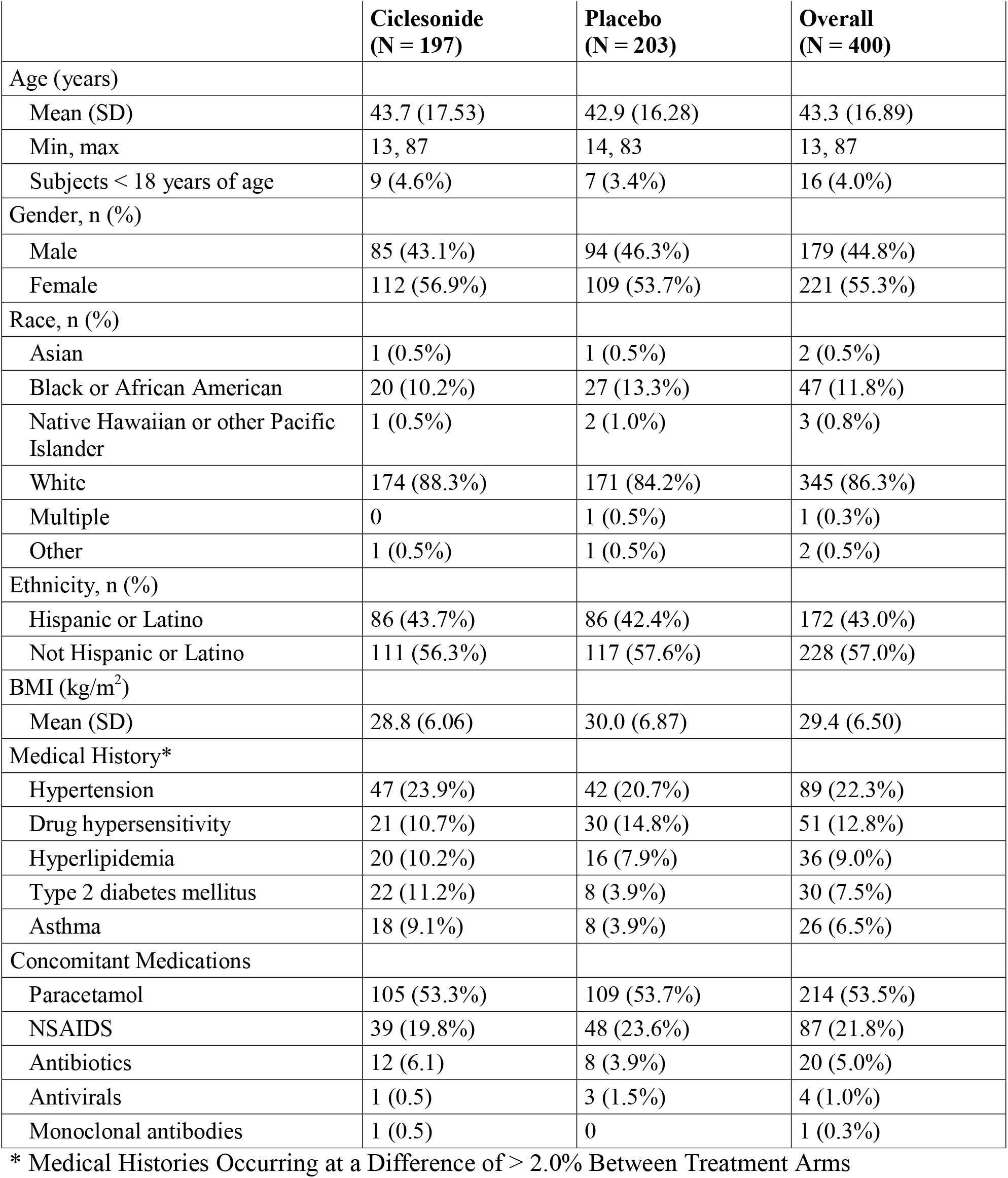
Participants’ Demographics, Medical Histories and Concomitant Medications.

**Figure 1.**
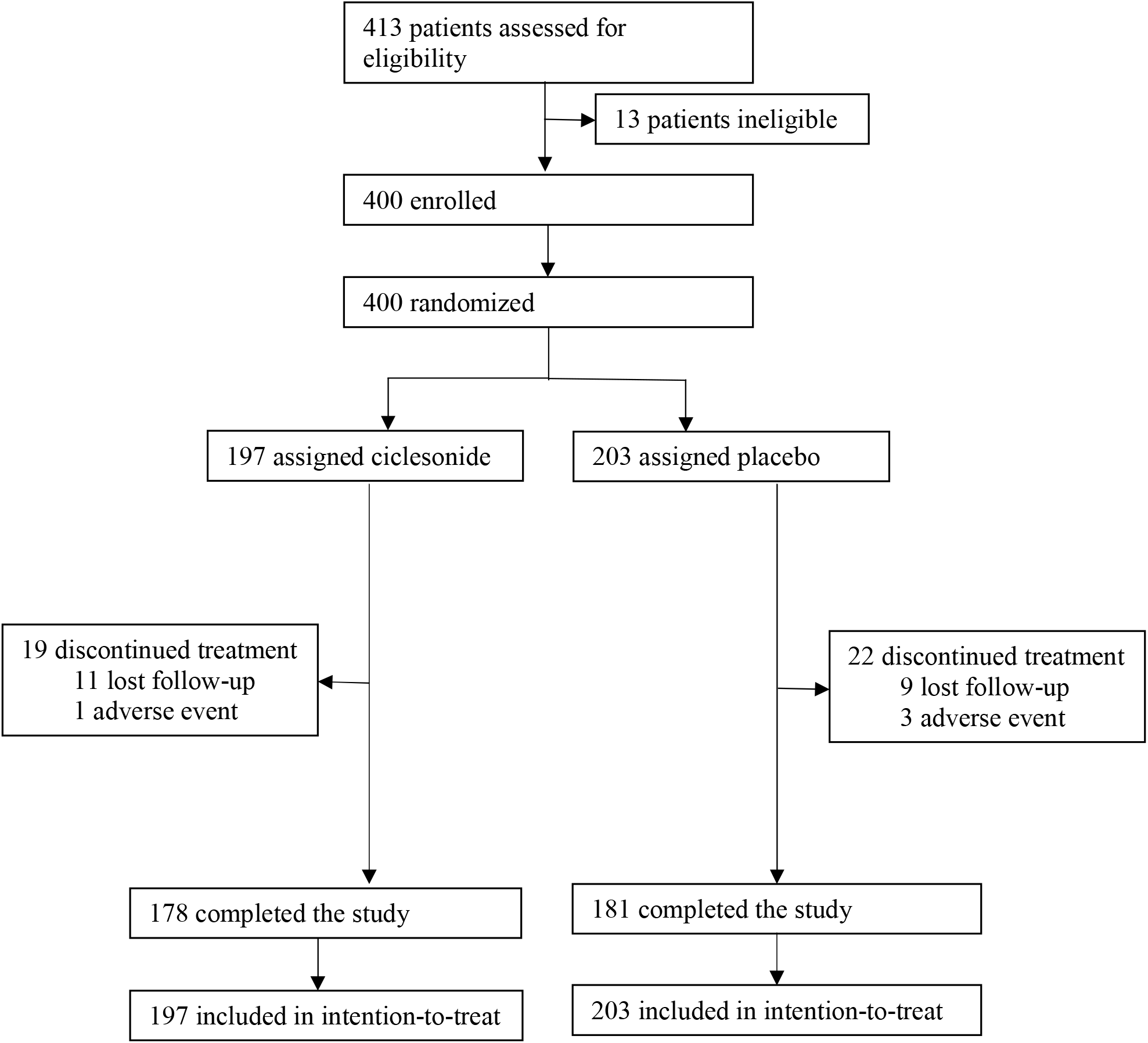
Trial profile.

The primary efficacy outcome was to assess whether treatment with ciclesonide MDI plus standard supportive care resulted in improved time to alleviation of COVID-19-related symptoms of cough, dyspnea, chills, feeling feverish, repeated shaking with chills, muscle pain, headache, sore throat, and new loss of taste or smell compared with placebo plus standard supportive care in non-hospitalized participants with symptomatic COVID-19 infection. In the ITT population, 139/197 (70.6%) participants in the ciclesonide arm and 129/203 (63.5%) participants in the placebo arm experienced alleviation of symptoms. Kaplan-Meier estimates of median time to alleviation of COVID-19-related symptoms were 19.0 days (95% CI: 14.0, 21.0) in the ciclesonide arm and 19.0 days (95% CI: 16.0, 23.0) in the placebo arm (Figure 2). The hazard ratio for the comparison of ciclesonide versus placebo based on a Cox proportional hazards regression model was 1.08 (95% CI: 0.84, 1.38).

**Figure 2.**
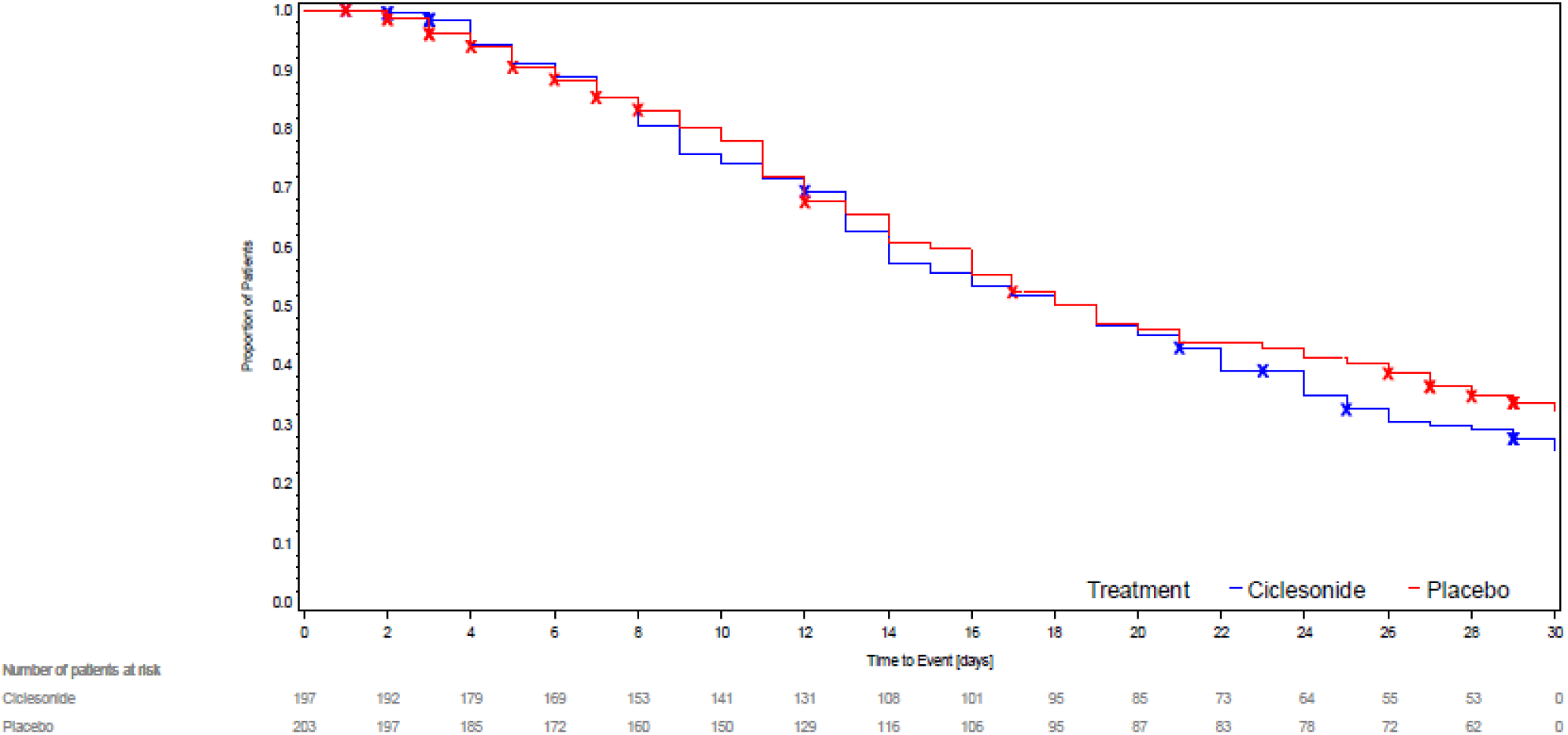
Kaplan-Meier Curve of Time to Alleviation of COVID-19-related Symptoms of Cough, Dyspnea, Chills, Feeling Feverish, Repeated Shaking with Chills, Muscle Pain, Headache, Sore Throat, and New Loss of Taste of Smell for a Continuous Period of ≥ 24 Hours (i.e., ≥ 3 AM/PM Assessments)

Participants receiving ciclesonide experienced fewer occurrences of emergency department visits or hospital admissions for reasons attributable to COVID-19 by Day 30 compared to those receiving placebo (1.0% vs 5.4%, odds ratio [OR] 0.18, 95% CI: 0.04 - 0.85, p=0.0301). No other secondary outcomes reached statistical significance. The most common symptoms reported on day 30 were cough (11.7% vs 12.3%, p=0.879), muscle pain (9.6% vs 8.9%, p=0.864) and dyspnea (10.2 vs 7.9, p=0.486).

Results of analysis of all secondary efficacy outcomes for the ITT population, including p-values and ORs (95% CI) for the comparison of ciclesonide versus placebo based on a logistic regression model adjusted for baseline covariates of sex, age, and baseline body mass index (BMI) are shown in Table 2. Sensitivity analyses for both the primary and secondary efficacy endpoints showed similar results as those from the corresponding primary analyses.

**Table 2.** Secondary Efficacy Outcomes.

| **Secondary Efficacy Endpoint** | **Ciclesonide**<br>**(N=197)** | **Placebo**<br>**(N=203)** | **Results** | **P-value** |
| --- | --- | --- | --- | --- |
| Percentage of participants with subsequent emergency department visit or hospital admission for reasons attributable to COVID-19 by Day 30 | 2 (1.0%) | 11 (5.4%) | Odds Ratio (ciclesonide vs placebo) (95% CI): 0.18 (0.04, 0.85) | 0.0301 |
| Percentage of participants with hospital admission or death by Day 30 | 3 (1.5%) | 7 (3.4%) | Odds Ratio (ciclesonide vs placebo) (95% CI): 0.45 (0.11, 1.84) | 0.2640 |
| All-cause mortality by Day 30 | 0 | 0 | N/A | N/A |
| COVID-19-related mortality by Day 30 | 0 | 0 | N/A | N/A |
| Percentage of participants with alleviation of COVID-19-related symptoms by Day 7 | 28 (14.2%) | 29 (14.3%) | Odds Ratio (ciclesonide vs placebo) (95% CI): 0.92 (0.51, 1.66) | 0.7865 |
| Percentage of participants with alleviation of COVID-19-related symptoms by Day 14 | 81 (41.1%) | 76 (37.4%) | Odds Ratio (ciclesonide vs placebo) (95% CI): 1.19 (0.78, 1.81) | 0.4283 |
| Percentage of participants with alleviation of COVID-19-related symptoms by Day 30 | 139 (70.6%) | 129 (63.5%) | Odds Ratio (ciclesonide vs placebo) (95% CI): 1.28 (0.84, 1.97) | 0.2534 |
| Hospital admission or death | 3 (1.5%) | 7 (3.4%) | Hazard Ratio* (ciclesonide vs placebo) (95% CI): 0.39 (0.09, 1.65) | 0.2008 |
| Change from baseline in oxygen saturation levels at Day 7 AM |  |  | Between-treatment difference in change from baseline (ciclesonide – placebo) (95% CI): -0.08 (-0.38, 0.22) | 0.6065 |
| Change from baseline in oxygen saturation levels at Day 7 PM |  |  | Between-treatment difference in change from baseline (ciclesonide – placebo) (95% CI): -0.10 (-0.36, 0.16) | 0.4655 |
| Change from baseline in viral load at Day 30 |  |  | Between-treatment difference in change from baseline (ciclesonide – placebo) (95% CI): -149.32 (-511.58, 212.93) | 0.4178 |

\*Hazard Ratio for time to hospital admission or death

Adverse events were reported by 22 (11.2%) participants in the ciclesonide arm and 29 (14.3%) participants in the placebo arm. Most adverse events were mild to moderate in severity. Oral candidiasis was reported in 1 (0.5%) participant in each of the study arms. Dry mouth was reported in 3 (1.5%) participants in the ciclesonide arm and 1 (0.5%) participant in the placebo arm. Distinct from the participants’ eDiaries reporting, headache was reported as an adverse event in 1 (0.5%) participant in the ciclesonide arm and 4 (0.5%) participants in the placebo arm. Discontinuation of study treatment due to 1 or more adverse events occurred in 3 (1.5%) participants in the ciclesonide arm and 7 (3.4%) participants in the placebo arm. One (0.5%) participant in each arm reported a headache which were judged to be adverse events in the opinion of the site principal investigators and were subsequently discontinued from the protocol. All other discontinuations due to adverse events were the result of hospitalizations, which was a predetermined discontinuation criterion. One (0.5%) participant in the ciclesonide was hospitalized for treatment of an animal bite and 1 (0.5%) participant in placebo arm was hospitalized for treatment of a bowel obstruction, neither were immediately discontinued. No participants died during the trial (Table 3).

**Table 3.** Overall Summary of Adverse Events.

| **Participants with at least 1 treatment-emergent** | **Ciclesonide (N = 197)** | **Placebo (N = 203)** |
| --- | --- | --- |
| Adverse event | 22 (11.2%) | 29 (14.3%) |
| Respiratory, thoracic and mediastinal disorders | 6 (3.0%) | 9 (4.4%) |
| Infections and infestations | 6 (3.0%) | 7 (3.4%) |
| Gastrointestinal disorders | 3 (1.5%) | 6 (3.0%) |
| Nervous system disorders | 2 (1.0%) | 6 (3.0%) |
| General disorders and administration site conditions | 1 (0.5%) | 2 (1.0%) |
| Renal and urinary disorders | 2 (1.0%) | 0 |
| Injury, poisoning and procedural complications | 2 (1.0%) | 0 |
| Musculoskeletal and connective tissue disorders | 2 (1.0%) | 0 |
| Cardiac disorders | 0 | 2 (1.0%) |
| Psychiatric disorders | 0 | 2 (1.0%) |
| Skin and subcutaneous tissue disorder | 1 (0.5%) | 1 (0.5%) |
| Metabolism and nutrition disorders | 1 (0.5%) | 0 |
| Investigations | 0 | 1 (0.5%) |
| Adverse event related to investigational product | 7 (3.6%) | 5 (2.5%) |
| Adverse event leading to study treatment discontinuation | 3 (1.5%) | 7 (3.4%) |
| Adverse event leading to death | 0 | 0 |

\* Subjects were followed for 60 days for adverse events, inclusive of 30-day treatment period.

## Discussion

No statistically significant difference was observed between participants treated with ciclesonide versus placebo for the primary efficacy endpoint.

These composite efficacy outcomes were based on resolution of all COVID-19 symptoms. It is not uncommon for COVID-19 patients to continue to have one or more mild lingering symptoms as they convalesce. Loss of smell, in particular, is a frequently reported symptom of COVID-19, which can be present for 3 weeks or more in many patients^11^. The endpoint of complete symptom recovery across all COVID-19 symptoms may have masked a significant population who were able to safely return to their baseline activities and were no longer at high risk for transmission, but who still had not entirely returned to their baseline.

In this study, participants treated with ciclesonide were less likely to have a subsequent emergency department visit or hospital admission for reasons attributable to COVID-19 by Day 30 (1.0% vs 5.4%, p = 0.03). The sensitivity analysis conducted on the PP population showed similar results. While secondary outcomes are considered exploratory, it is noteworthy that this was originally the primary outcome included in the initial study registration. This outcome may be more relevant to the patients and the health care systems than complete resolution of symptoms. Inhaled steroids may represent a relatively low-cost intervention to prevent emergency department visits or hospital admissions due to COVID-19. In this study, the number need to treat prevent emergency department visits or hospital admissions due to COVID-19 was 23.

Two recent open-label, randomized controlled trials of inhaled budesonide in the treatment of patients with COVID-19, demonstrated a decreased need for COVID-19-related urgent medical care^12^, and hospitalizations or death^13^, which is consistent with this study’s findings. Unlike this study’s findings, the studies of budesonide also demonstrated a decrease in time to symptom resolution^12, 13^.

Among the various completed or ongoing trials (NCT04193878, NCT04330586, NCT04331054, NCT04331470, NCT04355637, NCT04356495, NCT04377711, NCT04381364, NCT04416399, ISRCTN86534580) of inhaled corticosteroids for the treatment of COVID-19, few utilize a double blinded design. A double blinded design, like the one used in this study, is especially critical when relying on participant dependent endpoints such as self-reported symptoms and the decision to seek emergency department care.

Further studies of the efficacy of inhaled steroids among populations of older patients and patients with known risk factors are needed to explore the efficacy of inhaled steroids among patients at higher risk for severe diseased progression, hospitalization, and death from COVID-19.

This study followed participants for 60 days. As the pandemic continues, a growing population of patients with long-term COVID-19 symptoms or post-acute sequelae of SARS-CoV-2 beyond 12 weeks has emerged^14^. Longer-term studies are needed to better understand factors that may influence this growing subset of patients.

Ciclesonide did not achieve the primary efficacy endpoint of time to alleviation of all COVID-19-related symptoms. Future studies of inhaled steroids are needed to explore their efficacy in patients with high risk for disease progression and in reducing the incidence of long-term COVID-19 symptoms or post-acute sequelae of SARS-CoV-2.

## Supporting information

CONSORT Checklist

## Data Availability

Individual participant data that underlie the results reported in this article will be available after de-identification. The study protocol will also be available immediately following publication, ending 36 months following article publication.

## Identifying Data

NCT04377711

A Study on the Safety and Efficacy of Ciclesonide in the Treatment of Non-hospitalized COVID-19 Patients

## Contributors

MB and DK contributed to the study design. BC, RV, YGR, CM, and WP were site investors. DK did the statistical analysis. BC wrote the first draft of the report with input from MB, RV, YGR, CM, and WP, as well as medical writers (SHR and YJS). All authors contributed to manuscript revisions, had access to the data and had final responsibility for the decision to submit for publication. BC and DK have accessed and verified the data.

## Acknowledgements

The authors thank Skyler Hime-Rupard, MS and Yolaine Jeune-Smith, PhD of Cardinal Health Specialty Solutions, Dublin, OH for providing medical writing support, which was funded by Covis Pharma GmbH.

